# Reevaluation of Seroprevalence using a Semi-quantitative Anti-spike IgG in Health Care workers at an Academic Medical Center in Boston, Massachusetts

**DOI:** 10.1101/2022.01.20.22269543

**Authors:** Manisha Cole, Elizabeth R. Duffy, Jordyn N Osterland, Susan Gawel, Lei Ye, Kyle de la Cena, Elizabeth J. Ragan, Sarah E. Weber, Elissa M- Schechter-Perkins, Tara C. Bouton, Karen R. Jacobson, Chris Andry, Yachana Kataria

## Abstract

**Background:** Accurate measurement of antibodies is a necessary tool for assessing exposure to SARS-CoV-2 and facilitating understanding of the role of antibodies in immunity. Most assays are qualitative in nature and employ a threshold to determine presence of antibodies. Semi-quantitative assays are now available. Here we evaluate the semi-quantitative SARS-CoV-2 IgG II (anti-spike (S)) assay. We aim to reassess the seroprevalence using anti-S assay and subsequently compare it to the previously measured IgG (anti-nucleoprotein (N)) in health care workers at an academic medical center in Boston.

**Methods:** 1743 serum samples from HCWs at Boston Medical Center were analyzed for SARS-CoV-2 anti-S IgG and IgM using the Abbott SARS-CoV-2 IgG II and□Abbott AdviseDx□SARS-CoV-2 IgM assay, respectively. Precision, linearity, positive and negative concordance with prior RT-PCR test were evaluated for anti-S IgG. Seroprevalence and its association with demographics variables was also assessed.

**Results:** Linearity and precision results were clinically acceptable. The positive and negative concordance for anti-S IgG with RT-PCR was 88.2% (95% CI: 79.4% - 94.2%) and 97.43% (95% CI: 95.2% - 98.8%), respectively. Overall, 126 (7.2%) of 1,743 participants were positive by anti-S IgG. Among the 1302 participants with no prior RT-PCR, 40 (3.1%) were positive for anti-S IgG antibody. The original agreement in this population with the qualitative, anti-N IgG assay was 70.6%. Upon optimizing the threshold from 1.4 to 0.49 S/CO of the anti-N IgG assay, the positive agreement of the assay increases to 84.7%.

**Conclusion:** The anti-S IgG assay demonstrated reproducible and reliable measurements. This study highlights the presence of asymptomatic transmission among individuals with no prior history of positive RT-PCR. It also highlights the need for optimizing thresholds of the qualitative SARS-CoV-2 IgG assay for better agreement between assays by the same vendor.

## Introduction

Coronavirus disease 2019 (COVID-19) is the disease caused by the novel severe acute respiratory syndrome coronavirus 2 (SARS-CoV-2) that causes fever, cough, shortness of breath, and fatigue that can quickly become life threatening (1-3). SARS-CoV-2 RNA testing alone, in the absence of serological testing, is not sufficient to assess population-level viral transmission and pathogen exposure and the public health burden of the pandemic (4). The transient nature of RNA testing makes it an inaccurate metric to assess viral transmission at a population level. Accurate measurement of antibodies can facilitate understanding the role of antibodies in immunity elicited by both natural and vaccine response (5, 6). Ongoing studies are investigating the durability of antibodies against SARS-CoV-2 and examining what levels confer protective immunity against severe disease and/or reinfection (6-9).

There has been a rapid increase in serological assay availability in the United States via Emergency Use Authorization (EUA) from the Food and Drug Administration (FDA)(4, 10). These assays are designed to detect different immunoglobulin classes (IgM, IgG, IgA) or total antibodies to various epitopes of SARS-CoV-2. SAR-CoV-2 epitopes include the spike (S) or nucleocapsid (N) proteins. Most currently approved assays are qualitative in nature and employ a threshold to determine the presence of antibodies. Semi-quantitative assays are now available. To date, there are over 15 semi-quantitative assays approved for clinical testing in the United States including Roche Elecsys Anti-SARS-CoV-2 S, Phadia, Beckman Coulter, Inc. Access SARS-CoV-2 IgG II, AB EliA SARS-CoV-2-Sp1 IgG Test Phadia, Dimension EXL SARSCoV2 IgG, and Quanterix Simoa Semi-Quantitative SARS-CoV-2 IgG Antibody Test among others (11). However, it remains unclear if the pre-defined threshold for current qualitative and semi-quantitative assays is appropriately set (4, 12). It is also unclear whether these SARS-CoV-2 serological assays are comparable which prohibits assessment of durability and correlation with protection to help inform public health policies (4, 13).

We previously assessed seroprevalence among health care workers at an (11)(11)(11)academic medical center in Boston, Massachusetts, USA using the qualitative assay SARS-CoV-2 IgG (anti-nucleoprotein (N)) assay (11). Seroprevalence studies can assist in detecting asymptomatic and symptomatic infection and provide a cumulative prevalence estimate. Abbott Laboratories has now been issued an EUA for its anti-spike quantitative SARS-CoV-2 IgG II (anti-S IgG) and IgM (anti-S IgM) assay (6). In this study, we aim to reevaluate the seroprevalence in health care worker (HCW) population using the anti-S IgG assay and compare it to the anti-N IgG assay.

## Methods

### Study Design & Participants

Baseline samples were obtained from an ongoing longitudinal cohort assessing COVID-19 serological status among HCW’s at Boston Medical Center (BMC). In brief, participants were adult BMC employees who worked on campus during the first COVID-19 surge in Boston, MA (March 13th-May 31st, 2020). Baseline serum samples were obtained between July 13th-26th, 2020 (n= 1,743) and had been previously analyzed for anti-N IgG antibody status (Abbott Laboratories, Abbott Park, IL). Participants completed extensive questionnaire data on demographics (age, gender, race, ethnicity) between January 1^st^ – My 31^st^, 2020. Prior RT-PCR COVID-19 test results completed at BMC during the same period, were confirmed in the electronic medical record. This project was approved by the Institutional Review Board at Boston University Medical Center (BUMC).

For the current investigation, we analyzed serum samples for SARS-CoV-2 anti-S IgG & IgM (n = 1,743). These assays were performed by the clinical pathology laboratory at BMC on the Abbott Architect i2000 Instrument (Abbott Laboratories, Abbott Park, IL). Assays were run per the manufacturer’s instructions. For both assays, aliquots of serum samples (∼500 ul) were thawed for either 24 hours at 4°C or 2 hours at room temperature prior to being analyzed.

### SARS-CoV-2 IgM Assay (Qualitative)

The□AdviseDx□SARS-CoV-2 IgM (anti-S IgM) assay is <u>a chemiluminescent microparticle immunoassay (CMIA) for the qualitative detection of IgM antibody in human serum against the</u>S<u>ARS-CoV-2 spike protein.□□</u>

In this automated assays, participant serum, paramagnetic particles coated with SARS-CoV-2 antigen, and an assay diluent are incubated together during which the antibodies present in the serum sample bind to the antigen. The resulting luminescence will be read and resulted as a relative light unit (RLU). Both assays rely on an assay-specific calibrator to report a ratio of specimen RLU to calibrator RLU. Interpretation of positivity is determined by an index value above a predefined threshold (8, 9).

IgM samples were interpreted as positive (index value ≥ 1.00) or negative (index value < 1 .00). Both qualitative and quantitative results were used in the analysis.

### Anti-S IgG Assay (Semi-Quantitative)

Aliquots of serum samples (∼500 ul) were thawed for either 24 hours at 4°C or 2 hours at room temperature prior to being analyzed.

The Abbott ARCHITECT SARS-CoV-2 IgG II assay is a semi-quantitative assay that detects IgG antibodies to the receptor binding domain (RBD) of the S1 subunit of the spike protein of SARS-CoV-2 in human plasma. It is a two-step indirect CMIA.

The assay utilizes magnetic microparticles coated with RBD recombinant protein. The mixture is incubated and then washed prior to the addition of an anti-human-IgG antibody conjugated to acridinium. The recorder molecule is incubated prior to a second wash and followed by addition of a triggering solution that generates luminescence. The light is captured and counted to give an RLU. The RLU’s are read of a six-point calibration curve stored on the instrument generating an AU/mL (arbitrary units/milliliter) value. The assay is standardized to a monoclonal antibody concentration (14). Samples were interpreted as positive when >= 50.0 AU/mL or negative when <50.0 AU/mL. Both qualitative and quantitative results were used in the analysis.

### Precision SARS-CoV-2 IgM & Anti-S IgG

Precision studies were performed on quality control (QC) material as supplied by Abbott Diagnostics. IgM has two levels of QC: negative and positive. Anti-S IgG has three levels of QC: negative, low positive, and high positive. Intra-day precision was assessed by analyzing 20 replicates of each level of QC on the same day. Inter-day precision was assessed by analyzing QC for 20 days.

### Linearity Anti-S IgG Studies

The analytical measuring range of anti-S IgG is 22.0 - 25,000 AU/mL. The suggested manufacturer dilution is 1:2, extending the measuring upper limit to 50,000 AU/mL. Dilution studies were carried out using two participants that had elevated serum samples. Although the EUA approved upper linearity is 25,000 AU/mL, reagent was received prior to EUA approval and was reported to be 50,000 AU/mL at the time of these analyses. These samples were diluted serially with negative control. A total of 7 levels were tested in triplicates on the Abbott Architect i2000 instrument.

### Statistical Analysis

Questionnaire data were collected and managed in REDCap electronic data capture tools hosted at Boston University, CTSI 1UL1TR001430 (15, 16). Imprecision was assessed by measuring mean, standard deviation (SD), and coefficient of variation (CV) for inter and intra-day precision. Anti-S IgG data were log transformed (base 2) for analysis and ease of visualization.

Categorical data are presented as counts. We tested the association between anti-S IgG with sex, race, and gender using a Fisher’s Exact Test analysis. Missing data of less than 5% was excluded from analysis.

Positive and negative concordance of anti-S IgG was calculated using molecular testing (RT-PCR) as the gold standard among participants with a prior RT-PCR test result. Comparison between assays was assessed by Mcnemar’s chi-squared test. Receiver operating characteristics (ROC) curve was used to assess assay thresholds and define trade-offs in assay sensitivity and specificity using to RT-PCR test as the gold standard.

A P-value of < 0.05 was considered statistically significant, and all tests were two-sided. Analyses were performed in R Version R-3.5.3 (R Foundation for Statistical Computing). Select figures were produced by GraphPad Prism version (9.0.2) by GraphPad Software (San Diego, USA).

## Results

### Precision

IgM negative and positive control exhibited an intra-day CV of 16.5%, and 2.2%, respectively. IgM negative and positive control exhibited an inter-day CV of 21.3%, and 1.9%, respectively **(Supplementary Table 1)**.

Anti-S IgG negative, low positive, and high positive control exhibited an intra-day CV of 23.9%, 3.3%, and 2.9%, respectively. Anti-S IgG negative, low positive, and high positive control exhibited an inter-day CV of 16.6%, 2.4%, and 2.4%, respectively **(Supplementary Table 1)**.

### Anti-S IgG Linearity

Linearity data is shown in **Figure 1. Figure 1a** displays a participant result with greater than the analytical measuring range (46,820 AU/mL) and shows an acceptable linear response with a r-squared value of 0.99. Similarly, **Figure 1b** displays a participant result that had an original anti-S IgG level of 11,711.60 AU/mL with acceptable linearity with a r-squared value of 0.99, though not at the upper end of the analytical measuring range.

**Figure 1.**
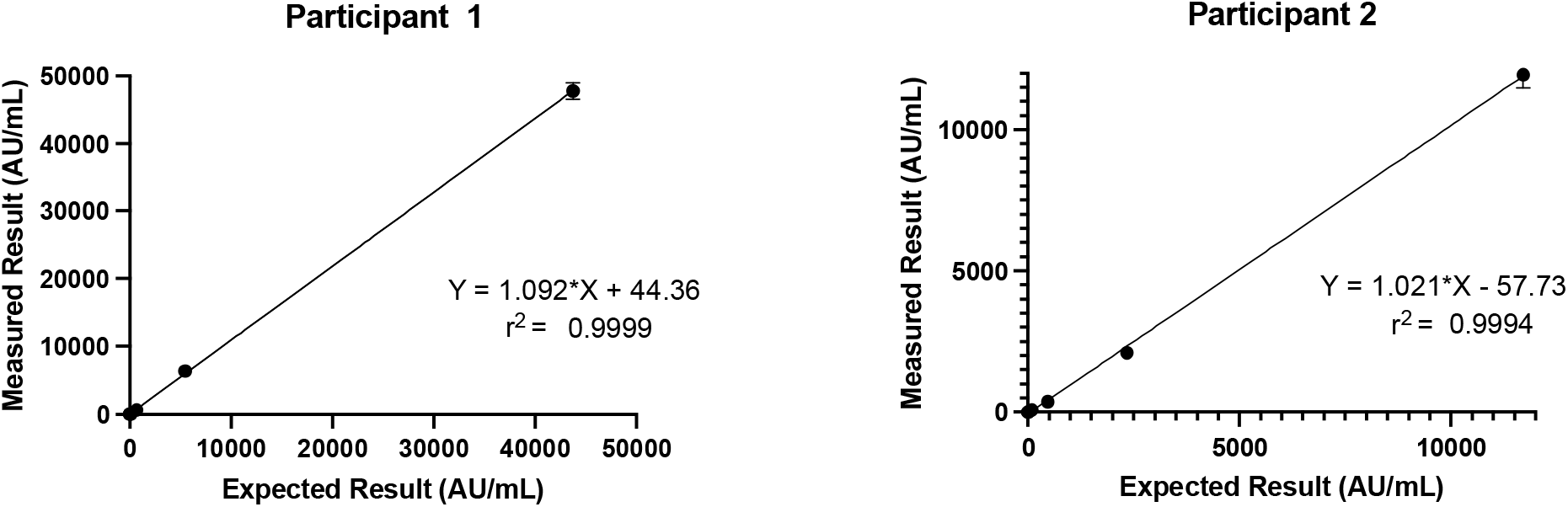
Linearity of Anti-S IgG

The data suggests that the difference between expected and measured is within the 95% CI. Significant percentage differences are observed on the lower end of the analytical measuring range **(Supplementary Figure 1)**.

### Seroprevalence

**Table 1** shows demographics in relation to anti-S IgG status (n = 1,743). Overall, 126 of 1,743 (7.23%) participants were anti-S IgG positive. Participants who were female, Hispanic, or Black were more likely to be seropositive, but these findings did not reach statistical significance. Obese participants were two times more likely to be seropositive for anti-S IgG [RR: 2.04 (95% CI: 1.40, 2.99)]. Among all the participants, anti-S IgG levels were not associated with age, race, or gender **(Figure 2a-c)**.

**Table 1.** Population Demographics by Anti-S IgG quant

| Anti-S IgG Status |  |  |  |  |  |
| --- | --- | --- | --- | --- | --- |
| Variable | Total<br>n (column %) | Positive<br>n (row %) | Negative<br>n (row %) | p-value<br>(All Data) | RR (95% CI)<br>(All Data) |
| Total | 1743 (100.00) | 126 (7.23) | 1617 (92.77) |  |  |
| Sex |  |  |  | 0.31 |  |
| Female | 1306 (74.93) | 102 (7.81) | 1204 (92.19) |  | Referent |
| Male | 431 (24.73) | 24 (5.57) | 407 (94.43) |  | 0.71 (0.46, 1.10) |
| Nonbinary/3rd Gender | 3 (0.17) | 0 (0.00) | 3 (100.00) |  | 0.00 (N/A)** |
| Average Age (years) (SD) | 41.16 (12.39) | 41.00 (12.21) | 41.17 (12.41) | 0.53 |  |
| BMI |  |  |  | p < 0.001* |  |
| Underweight (<18.5 kg/m²) | 34 (1.95) | 0 (0.00) | 34 (100.00) |  | 0.00 (N/A)** |
| Normal (18.5-24.9 kg/m²) | 844 (48.42) | 53 (6.28) | 791 (93.72) |  | Referent |
| Overweight (25.0-29.9 kg/m²) | 528 (30.29) | 29 (5.49) | 499 (94.51) |  | 0.87 (0.56, 1.36) |
| Obese (>29.9 kg/m²) | 335 (19.22) | 43 (12.84) | 292 (87.16) |  | 2.04 (1.40, 2.99) |
| Hispanic/LatinX |  |  |  | 0.10 |  |
| Yes | 143 (8.20) | 15 (10.49) | 128 (89.51) |  | Referent |
| No | 1594 (91.45) | 109 (6.84) | 1485 (93.16) |  | 0.65 (0.39, 1.09) |
| Race |  |  |  | 0.52 |  |
| White | 1299 (74.53) | 87 (6.70) | 1212 (93.30) |  | Referent |
| Asian | 160 (9.18) | 10 (6.25) | 150 (93.75) |  | 0.93 (0.50, 1.76) |
| Black | 141 (8.09) | 13 (9.22) | 128 (90.78) |  | 1.38 (0.79, 2.40) |
| Native American / Pacific Islander | 7 (0.40) | 0 (0.00) | 7 (100.00) |  | 0.00 (N/A)** |
| Other | 123 (7.06) | 12 (9.76) | 111 (90.24) |  | 1.46 (0.82, 2.59) |
| Smoking |  |  |  | 0.77 |  |
| No | 1671 (95.87) | 122 (7.30) | 1549 (92.70) |  | Referent |
| Yes | 46 (2.64) | 4 (8.70) | 42 (91.30) |  | 1.19 (0.46, 3.09) |

\* Statistically significant when p-value < 0.05 \*\*N/A: unable to divide by 0 Column percentages may not always add up to 100% due to missing data of <5.0%

**Figure 2.**
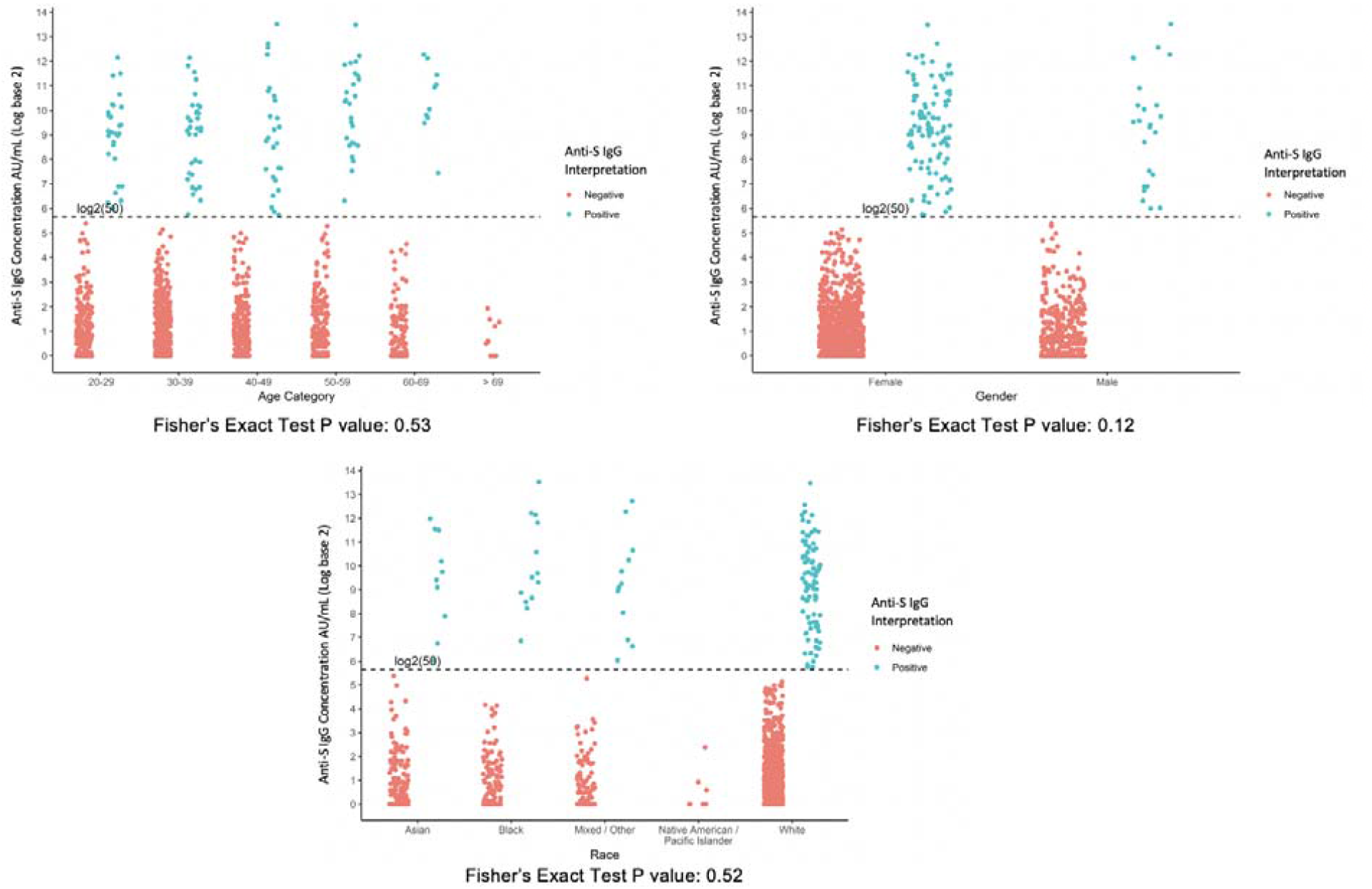
Anti- S IgG by demographics - a). Age b). Gender c). Race

441 of 1743 participants had a history of RT-PCR test. 47 of 85 RT-PCR confirmed positive individuals were also seropositive by IgM corresponding to a positive concordance of 55.3% (95% CI: 44.1% - 66.1%). 343 of 350 RT-PCR confirmed negative participants were seronegative by IgM corresponding to a negative concordance of 98.0% (95% CI: 95.9% - 99.2%) **(Supplementary Table 2)**. Of the remaining six participants with indeterminate RT-PCR results, two had detectable IgM antibody.

75 of the 85 RT-PCR confirmed positive participants were also seropositive by anti-S IgG corresponding to a positive concordance of 88.2% (95% CI: 79.4% - 94.2%). 341 of 350 RT-PCR confirmed negative participants were seronegative by anti-S IgG corresponding to a negative concordance of 97.43% (95% CI: 95.2% - 98.8%) **(Table 2)**. A total of 1302 participants had no prior RT-PCR test and of these participants, 40 (3.1%) were positive by anti-S IgG. Distribution of the anti-S IgG levels by RT-PCR test result is shown in **Figure 3**. Among the individuals with a prior RT-PCR, there is a statistically significant difference among the anti-S IgG levels by RT-PCR test result (p<0.001).

**Table 2.** Anti-S IgG vs. SARS-CoV-2 RT-PCR

|  |  | SARS-CoV-2 RT-PCR |  |  |  |
| --- | --- | --- | --- | --- | --- |
|  |  | Positive | Negative | IND* | Not Tested |
| Anti-S IgG | + | 75 | 9 | 2 | 40 |
|  | - | 10 | 341 | 4 | 1262 |

\*Indeterminate

**Figure 3.**
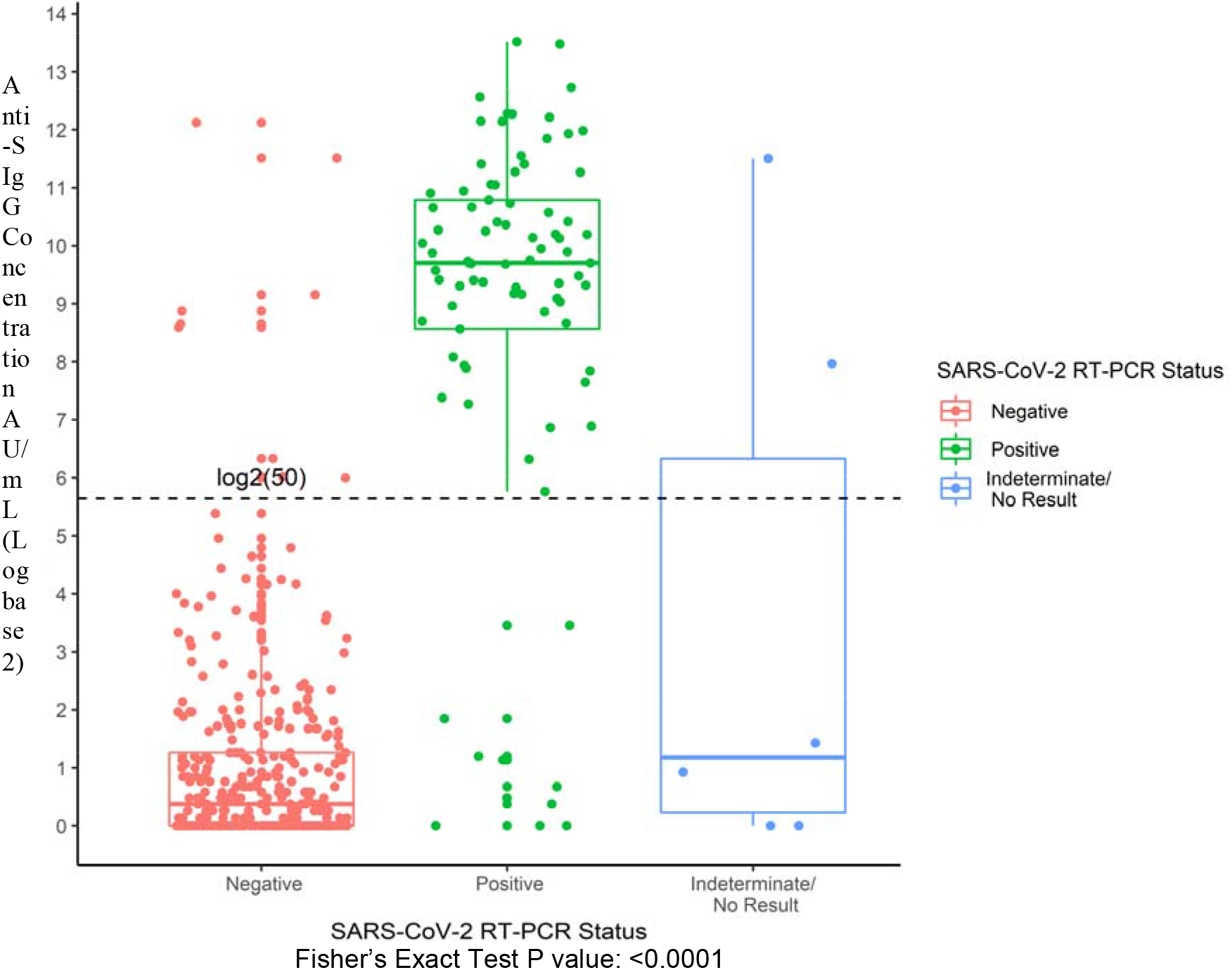
Anti-S IgG distribution by SARS-CoV-2 RT-PCR status

A total of 270 of the 441 individuals with a prior RT-PCR test had a recorded date of RT-PCR. **Figure 4** displays anti-S IgG level distribution since RT-PCR date. There was no statistical difference between anti-S levels in individuals less than 120 days after positive RT-PCR test result (p=0.16) **(Supplementary Figure 2)**. Anti-S IgG antibodies in RT-PCR confirmed individuals remained elevated over 120 days post infection.

**Figure 4.**
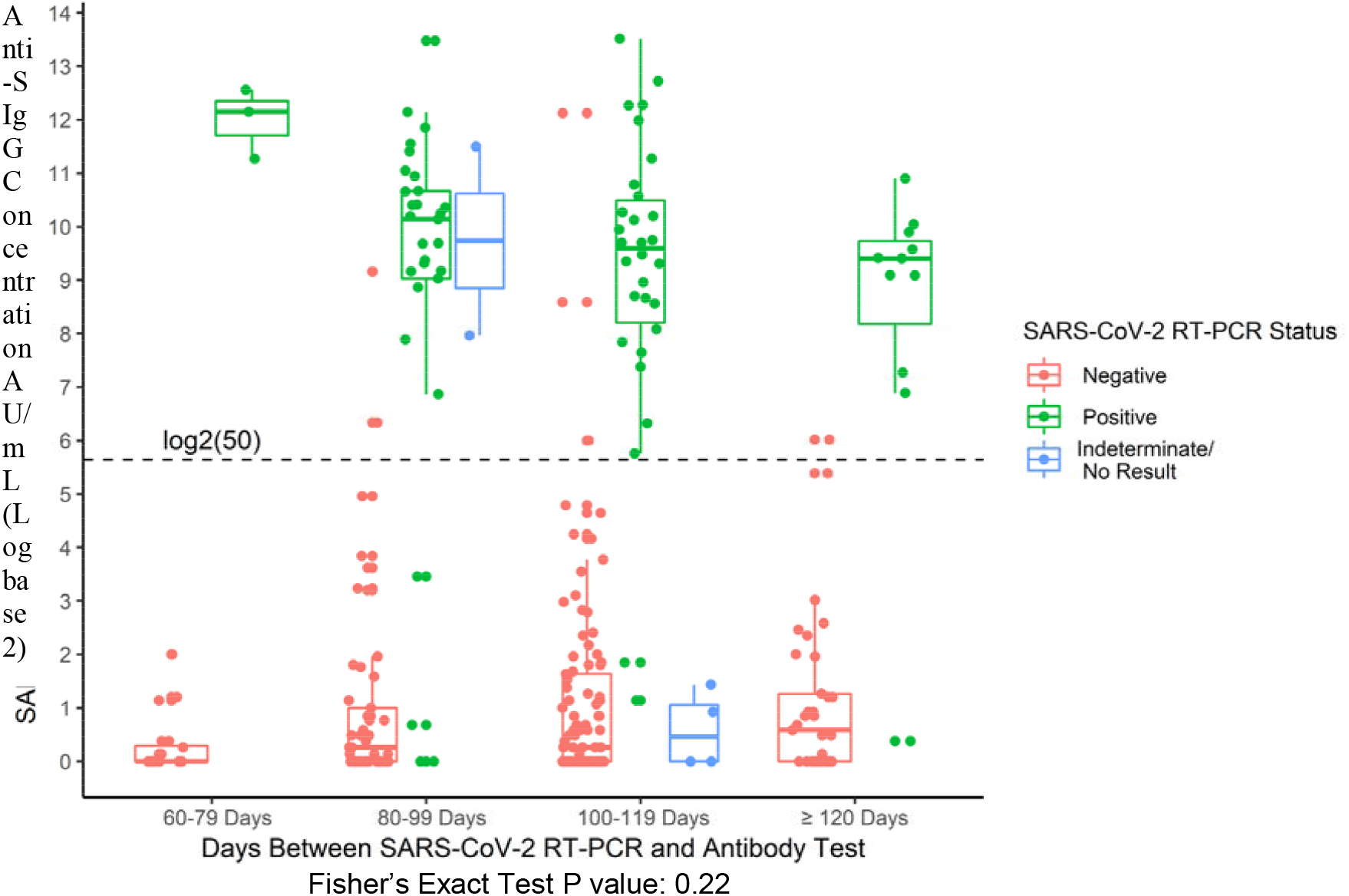
Anti-S IgG distribution days post SARS-CoV-2 RT-PCR result

### Agreement between Anti-N IgG vs. Anti-S IgG Status

Overall agreement between the two assays in this population was found to be 97.5% (95% CI: 96.7% - 98.2%). Of note, the positive agreement was 70.6% (95% CI: 61.8% - 78.4%), whereas the negative agreement was 99.6% (95% CI: 99.2% - 99.9%). These two tests were found to be statistically different (p<0.001).

A total of 37 participants had detectable anti-S IgG but were negative by anti-N IgG **(Table 3a)**. Of these, 11 participants had a positive RT-PCR, 3 participants had a negative RT-PCR, 1 participant had an indeterminate RT-PCR, and 22 were not tested. Of interest, 6 participants had detectable anti-N IgG results but were negative by anti-S IgG.

**Table 3.** Anti-N IgG vs. Anti-S IgG Agreement

|  |  |  |  |
| --- | --- | --- | --- |
| a. |  | Anti-S IgG |  |
|  |  | + | - |
| Anti-N IgG | + | 89 | 6 |
|  | - | 37 | 1611 |
| McNemar's Test Statistic with Yates' Continuity Correction: 20.93 P value: <0.0001 |  |  |  |
| b. |  | Anti-S IgG |  |
|  |  | + | - |
| Anti-N IgG | + | 109 | 21 |
|  | - | 17 | 1596 |
| McNemar's Test Statistic with Yates' Continuity Correction: 0.24 P value: <0.0001 |  |  |  |

### Assay Threshold Optimization

ROC curve analysis showed that the SARS-CoV-2 IgG qualitative had optimal sensitivity and specificity target e.g., adjusting threshold to 0.49 S/CO would result in a sensitivity of 84.7% and specificity of at least 96.9% (**Supplementary Figure 3**).

As such, changing the threshold of positivity for the anti-N IgG assay from 1.4 to 0.49 S/CO ratio increased the positive agreement between anti-N IgG & anti-S IgG from 70.6% to 86.5% (95% CI: 79.3% - 91.9%). However, it did not appreciably affect the negative agreement, from 99.6% to 98.7% (95% CI: 98.0% - 99.2%) (**Table 3b)**. Upon threshold modification, the two anti-N IgG vs. anti-S IgG were found to be statistically aligned.

## Discussion

This study reevaluates the seroprevalence in HCWs using a semi-quantitative anti-S IgG assay. It also compares the findings to the anti-N IgG in the same population. We report an overall seroprevalence of 7.2%. Among participants with a negative RT-PCR 2.5% of participants had detectable anti-S IgG antibody. We also observed a seroprevalence of 3.1% among participants with no prior RT-PCR. Our results demonstrate that the anti-S & anti-N IgG assays were statistically different. Upon lowering the anti-N IgG assay threshold, overall agreement between the two assays improved. Taken together, these results support the clinical utility of anti-S IgG assay and highlight the presence of COVID-19 infection among individuals with a negative or no prior RT-PCR.

The anti-S IgG shows acceptable intra- and inter-day precision. The precision for the negative control is expected to be high due to a lower numerical value and small variations observed artificially increase the CV. Our results depict acceptable linearity up to 50,000 AU/mL which exceeds upper limit stated in the package insert. It is currently unclear what titer levels confer SARS-CoV-2 immunity but our data suggest that such elevated levels are not rare among the participants.

Our results indicate less than ideal positive concordance for anti-S IgG vs. RT-PCR testing which is inconsistent with the manufacturer claims. Further investigation of the discordant samples (n=10) unveil that these participants did not have detectable anti-N IgG, or IgM antibody. These findings can possibly be explained by mild symptoms and/or disease. Literature suggests that the intensity and longevity of antibody response is associated with disease severity (17-19). The negative concordance of the SARS-CoV-2 assay was robust and consistent with the manufacturer claims.

Neither SARS-CoV-2 anti-N nor anti-S IgG assays find an association with sex, race, smoking status. However, the seroprevalence determined by anti-S IgG assay was higher (7.2%) relative to the anti-N IgG assay (5.5%) (8). The observed seroprevalence at a regional Boston area hospital was comparable to ours (20). Whereas others in the larger Boston region have reported a seroprevalence range between 14% to 25% (21). Seroprevalence can vary between sampling times and geographic regions. Among participants that had no prior RT-PCR test, 40 participants had detectable anti-S IgG antibody and 23 participants had detectable anti-N IgG antibody. As such, 17 additional individuals were identified to have IgG antibodies. This suggests higher sensitivity of the anti-S IgG assay which is corroborated by others (22). Alternatively, it may be attributable to a shorter half-life of anti-N IgG antibody (7).

These individuals pose a risk of asymptomatic viral transmission. Studies suggest that asymptomatic individuals are a source of transmission even after being vaccinated (23-25). Ultimately, RT-PCR testing is limited by the brief infectious window during which viral detection is possible. Detectable viral loads during infection via RT-PCR can vary by several factors such as patient medical history, immune response, and medications, whereas antibody measurements can detect exposure to virus for longer periods of time (26, 27). Our data suggests that participants in the study had elevated anti-S IgG for at least 120 days after RT-PCR tests. The longevity of detectable antibody remains unclear especially in less severe infections.

The qualitative nature of the anti-N IgG assay enabled us to optimize the pre-set threshold. Upon changing the anti-N IgG threshold, the assays exhibit better agreement with anti-S IgG. 20 of the 37 discordant participant samples became concordant. The remainder (14/17) discordant participants did not have a prior RT-PCR test, or a robust anti-N IgG response and most also (12/17) had a negative IgM. This suggests either a false positive anti-S IgG result or early detection of the seroconversion process in asymptomatic individuals. We have been following part of this cohort longitudinally and aim to determine if these individuals mount a permanent immune response over time. Furthermore, a study conducted by Public Health England also independently suggested the same optimized threshold to increase the sensitivity of the assay (28). Of interest, the same cut-off is currently used in Europe and provides external validation of our findings. Taken together, our data lend further support for optimizing the assay threshold to achieve better performance characteristics.

These differences between serological assays are also observed between vendors as well. In a head-to-head comparison of five semi-quantitative SARS-CoV-2 IgG assays found that the results from assays are not interchangeable despite good correlation to neutralizing antibody for some of them (10). This has been supported by others (9, 29, 30). Collectively, this highlights the need for harmonization between all serological assays. This is of paramount importance as it will enable us to draw meaningful conclusions about correlation of immunity and being better equipped to overcoming the SARS-CoV-2 pandemic.

The present study benefited from a large sample size. Certain limitations are acknowledged. The gold standard for determining protective antibody is a virus neutralization test which we were unable to compare to the anti-N & anti-S assay as it requires live pathogen and a biosafety level 3 laboratory. The study is limited by a cross sectional study design which may under- or overestimate the seroprevalence. We were unable to correlate days since RT-PCR result or viral load with quantitative antibody levels and it may provide insight about antibody kinetics (31). The samples were obtained from a single timepoint preventing characterization of antibody kinetics on performance characteristics; however, a subset of participants is being prospectively followed in three-month intervals which will enable future analysis. Lastly, the findings are not generalizable to the larger community and is limited to hospitals.

In conclusion, the Abbott SARS-CoV-2 anti-S IgG assay demonstrate acceptable performance characteristics. The study highlights the presence of infection among participants with no RT-PCR testing and among those with a negative RT-PCR test. It also highlights the need for optimizing thresholds of the qualitative SARS-CoV-2 IgG assay for better agreement between assays by the same vendor. Serological testing can aid in identify a better assessment of the public health burden.

## Data Availability

All data produced in the present study are available upon reasonable request to the corresponding author.

## Acknowledgements

We would like to acknowledge the clinical chemistry, phlebotomy, and central receiving staff in the Department of Laboratory Medicine and Pathology at Boston Medical Center for working with the research team to accomplish this study.

