## Supplemental Material for "Reevaluation of Seroprevalence using a Semi-quantitative Anti-spike IgG in Health Care workers at an Academic Medical Center in Boston, Massachusetts"

**Supplementary Figures and Tables:**

**Supplementary Table 1** – SARS-CoV-2 IgM and SARS-CoV-2 IgG II Precision

**Supplementary Figure 1** – Comparison of expected vs. measured SARS-CoV-2 IgG II using participant specimens

**Supplementary Table 2** - SARS-CoV- 2 IgM vs. SARS-CoV-2 RT-PCR

**Supplementary Table 3** - SARS-CoV- 2 IgG II and SARS-CoV- 2 IgG breakdown by SARS-CoV-2 RT-PCR status

**Supplementary Figure 2** – ROC analysis of SARS-CoV-2 IgG

**Supplementary Figure 3** - SARS-CoV-2 IgG II distribution at 120 days cut-off since SARS-CoV-2 RT-PCR

**Supplementary Table 1** – SARS-CoV-2 IgM and SARS-CoV-2 IgG II Precision

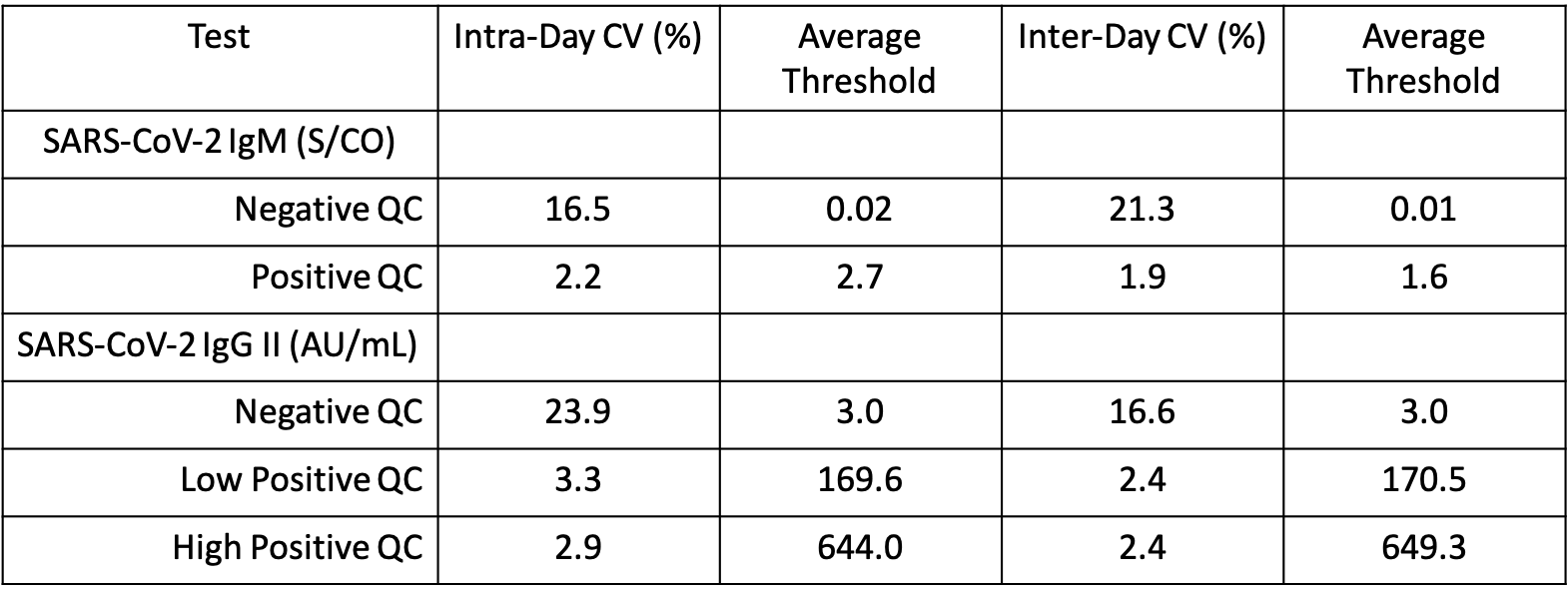

**Supplementary Figure 1** – Comparison of expected vs. measured SARS-CoV-2 IgG II using participant specimens

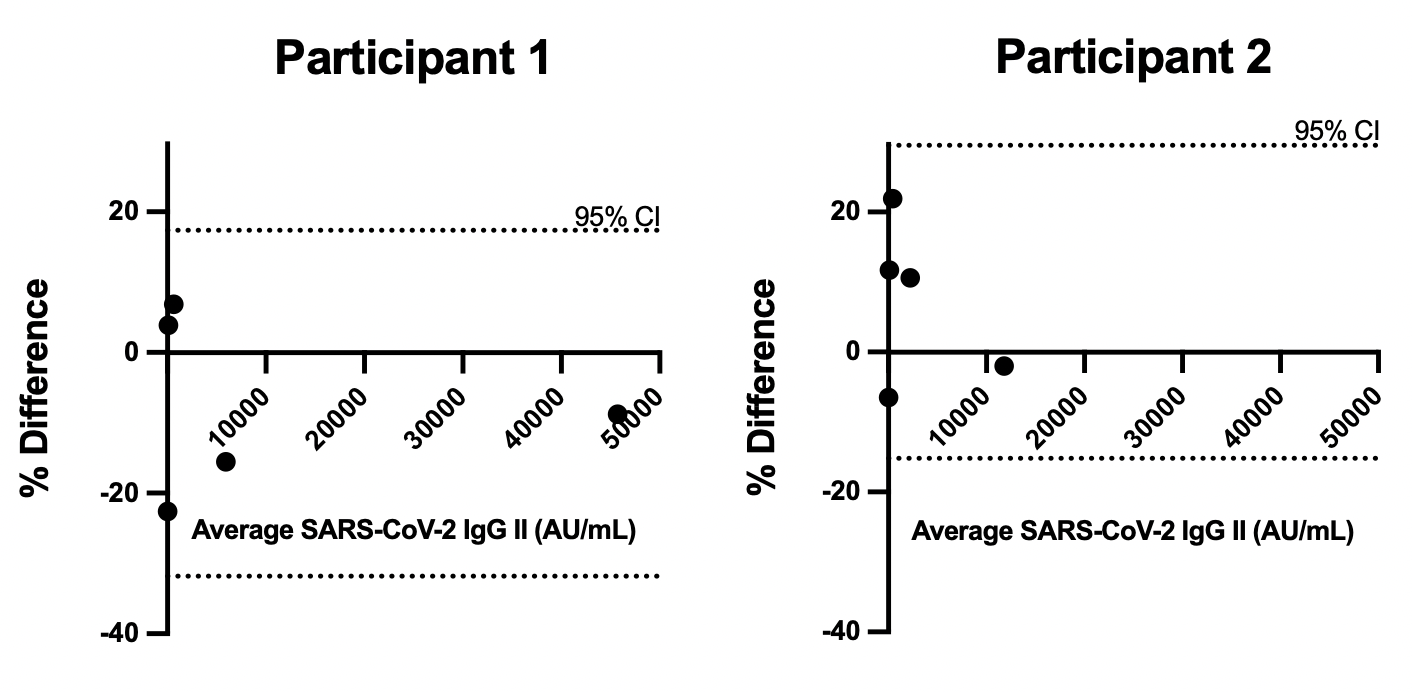

**Supplementary Table 2** - SARS-CoV- 2 IgM vs. SARS-CoV-2 RT-PCR

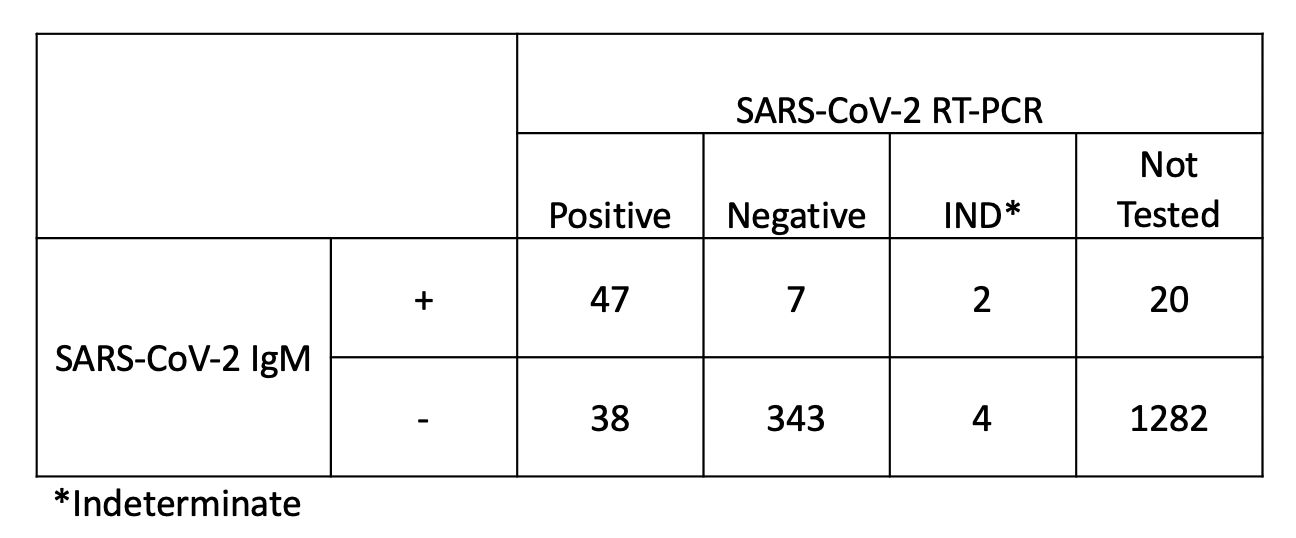

**Supplementary Table 3** - SARS-CoV- 2 IgG II and SARS-CoV- 2 IgG breakdown by SARS-CoV-2 RT-PCR status

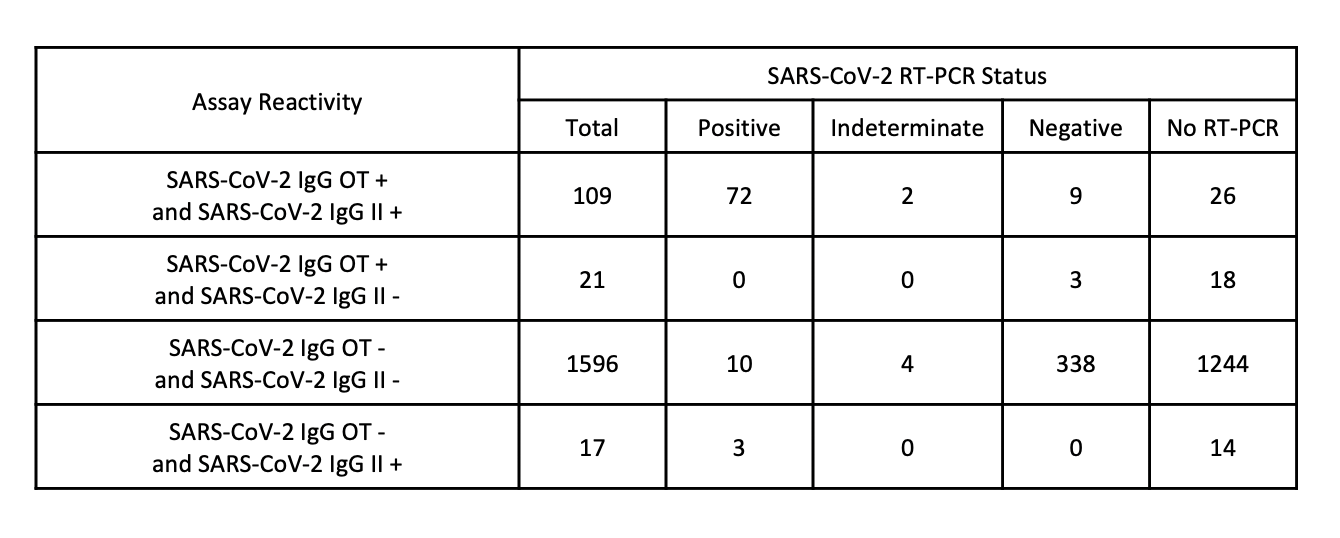
OT: Optimized Threshold

**Supplementary Figure 2** – ROC analysis of SARS-CoV-2 IgG

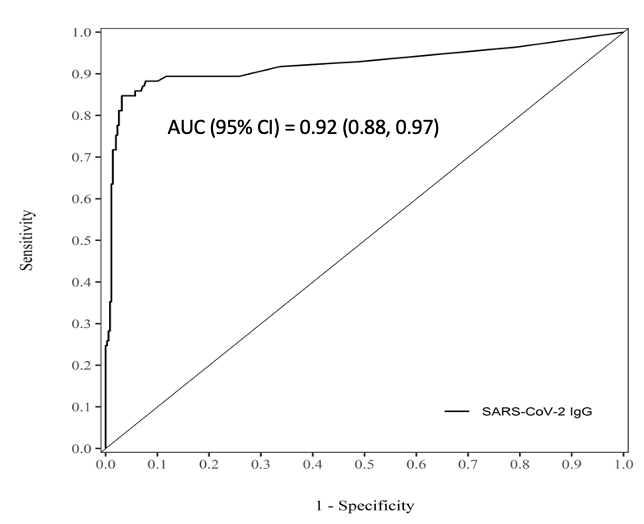

**Supplementary Figure 3** - SARS-CoV-2 IgG II distribution at 120 days cut-off since SARS-CoV-2 RT-PCR

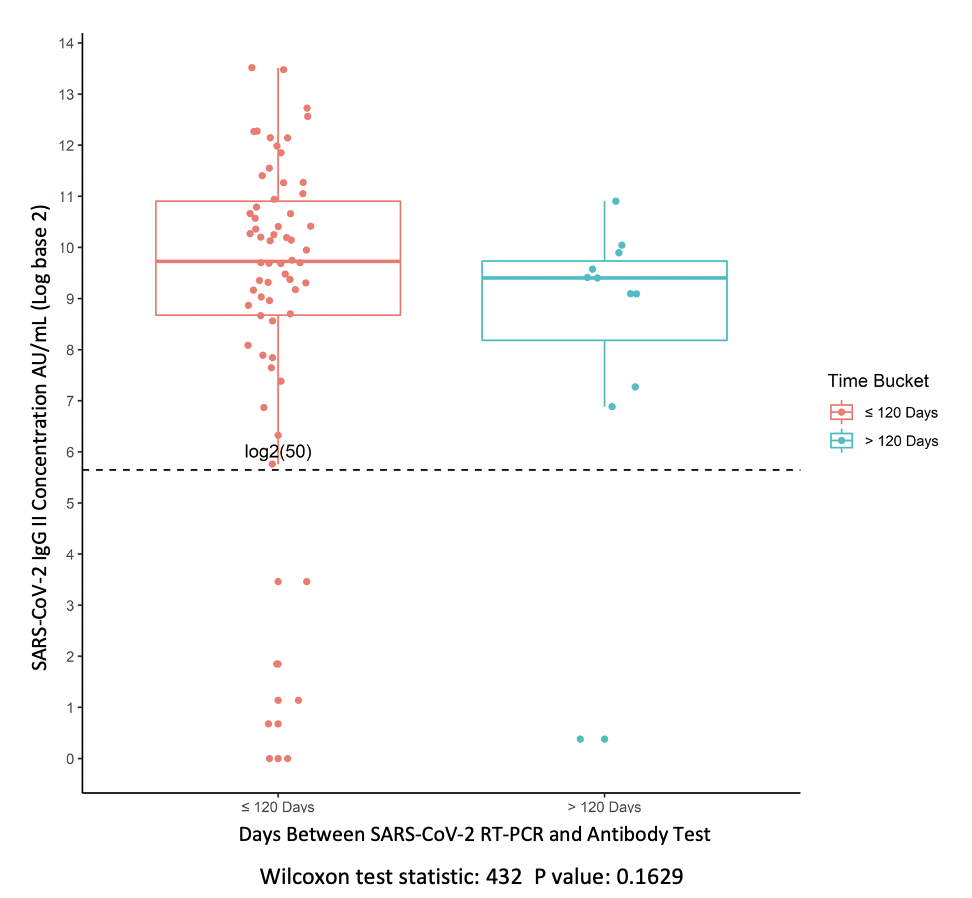
